# ASSESSING VACCINATION STRATEGIES FOR THE COVID-19 EPIDEMIC IN MINAS GERAIS (BRAZIL)

**DOI:** 10.1101/2021.04.19.21255731

**Authors:** Marcelo Marchesin, Mehran Sabeti

## Abstract

In this work we analyze the effectiveness of vaccination strategies for the COVID-19 epidemic in the Brazilian state of Minas Gerais. Firstly we study the effectiveness of general vaccination in the decreasing of the number of infected individuals using a traditional non structured SEIR model. Secondly we consider an age-structured SEIR model with 3 age groups (youngster, adult and elderly) and we analyze the current strategy in the Brazilian state of Minas Gerais, of focusing the vaccination on the elderly group. We conclude by showing this strategy to be mistaken and that a vaccination focusing on the age group of the adults would be much more efficient in decreasing the total number of infected individuals.

## 1. Introduction

Several cases of pneumonia of unknown etiology were detected in the Chinese city of Wuhan around the end of 2019. The World Health Organization ^1^was informed and reported that a novel corona virus (officially named COVID-19) was identified on January 7^*th*^ as the possible cause of such infections. An international alert was issued by WHO because of the imminent potential for the spreading of such an infectious disease worldwide. The disease quickly spread in all countries of the world with several different degrees of seriousness causing the WHO to declare it a pandemic on March 11^*th*^, due to the seriousness of the situation.

Although the WHO has constantly advised the international community for the need of social isolation and massive testing many countries, among which notably Brazil, have not taken the advices seriously enough and the number of infected individuals in the world has reached the incredible mark of 121, 164, 126 causing 2, 679, 841 deaths, leaving 97, 661, 975 recovered cases and 20, 822, 310 active cases by March 16^*th*^ a little bit over an year after the outbreak, according to the site Worldometers (see [16]).

Brazil has around 2.72% of the world population and yet has officially 11, 594, 204 cases of infected individuals which adds up to 9.57% of the total cases of the whole world. The official number of deaths in Brazil due to COVID-19 is 281, 626, indicating a lethality rate of around 2.43% while the world average lethality rate is around 2.21%. So Brazilian lethality rate is about 10% higher than the world average rate. Many reasons could be credited to such horrifying number from Brazil but in one word one can say the main cause of such a huge number of lives lost for COVID-19 was the Brazilian Government approach to the sanity crisis.

The Brazilian state of Minas Gerais, geographically located at the central part of Brazil with important and populous neighbor states as Rio de Janeiro and Sao Paulo has a dense web of road routes as well as an important aerial hub connecting several parts of the whole country. Minas Gerais has around 10% of the Brazilian population and a rich diversity of races, cultures, ages, genders and economic income distribution. All these reasons justifying us considering an analysis of the development of the spreading of the epidemic in the state of Minas Gerais as a good lab for the development of the disease nationwide.

After a very intensive effort of the international scientific community, the first samples of vaccines were presented to the world around the end of November of 2019. On January 20^*t*^*h* Brazil started it vaccination program yet at a very low rate of around 200, 000 individuals/day. The state of Minas Gerais started the vaccination program on January 25^*th*^ at an approximate rate of 20, 000 individuals/day. As it has being happening worldwide the elderly have been prioritized (besides the health agents) in this first stage of the vaccination program due to their clear vulnerability.

There are several forms of control measures that work by reducing the average amount of transmission between infectious and susceptible individuals. Which control strategies, and/or mix of strategies, are used will depend on the disease, hosts, and size of the epidemic [9]. One such tactic is continuous vaccination (permanent vaccination), another one is “pulse vaccination”, where instead of constantly vaccinating an extremely large proportion of all susceptible individuals, it is proposed to vaccinate a fraction of the entire susceptible population in a single pulse, which should be administered every T periods of time [13]. The importance of the use of vaccination, either continuous or pulse, as a means of protection against many diseases can be grounded on two counts: - The decrease in new cases of infectious diseases directly and indirectly, that is, both for those who have been vaccinated and for those who have not yet been. (This indirect protection produces an increase in the epidemic interval and therefore an increase also in the average age of infection.) - The low cost of the vaccine compared to treating the disease or not treating it at all. Theoretical results show that the “pulse vaccination” strategy can distinguish conventional leading strategies for disease eradication at relatively low vaccination values [1]. In contrast, conventional vaccination strategies are predicted to lead to epidemic control if the proportion of successfully vaccinated individuals is greater than a certain critical value.

The goal of this study is to assess through the analysis of different vaccination strategies the best one for the state of Minas Gerais. The reason behind our argument is that, assuming that the vaccination helps decreasing the rate of transmissibility of the disease, we claim that vaccination of elderly people, which are relatively socially isolated, will basically protect only the own vaccinated individual meanwhile vaccination of adults (or even youngster) would protect not only the vaccinated individual but several other individuals which are socially related to the vaccinated one leading to a quicker decrease in the total number of infected individuals.

We point out that even though Minas Gerais is considered for the scope of this paper, the techniques and tools used in this study can be easily adapted for any other city, state or country in the world. We point out that a more strict assess involving the strategies of vaccination of youngsters was neglected in this paper because most of the vaccines currently available in the world have not yet been tested in such an age group, thus practically no countries in the world are vaccinating individuals younger than 18 years old.

The paper is organized as follows: In section 2, the classical SEIR model with vaccination is presented. Based on the public official available data of the number of infected individuals we perform a fitting of the parameters and compare the curve of infected individuals given by the fitted model to the real number of cases. We further study the effectiveness of vaccination in the decreasing of total number of infected individuals using the classical SEIR model. In section 3, the age structured SEIR model with vaccination is presented. Due to the fact that the beginning of the vaccination program in Minas Gerais happened on January 25^*th*^, 2021 we have chosen to study the model considering the initial conditions to be the ones related to January 1^*st*^ of 2021. In section 4, after the parametric fitting for the age structured SEIR model is done, we use such a model to project the total number of infected individuals considering different strategies of vaccination depending on the parameters *ν*_1_, *ν*_2_ and *ν*_3_ which represent the rates of vaccination in the 3 analyzed age groups. The conclusion is presented in section 5.

## 2. The Classical SEIR model

The classical SEIR model with vaccination and without vital dynamics is given by

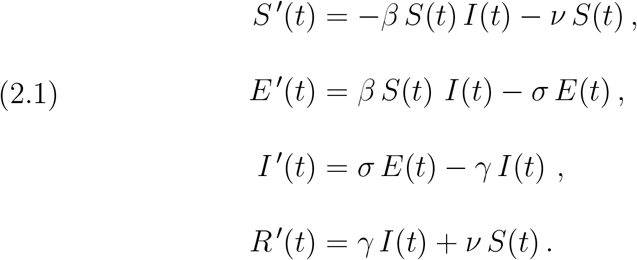

The above parameters are all non-negative, *β* represents the transmission rate, *σ* is the rate at which exposed individuals become infected, *γ* is the recovery rate and *ν* represents the rate of vaccination.

As we have already mentioned we shall consider the development of the epidemic from day one (i.e., January 1^*st*^, 2021) on. For this we consider the total number of infected individuals in Minas Gerais 546, 884 (see [12]). The parameters *β, σ* and *γ* can be adjusted so that the SEIR curve fits well the real data. To achieve that, the difference between the SEIR curve and the real data values for the number of infected individuals is minimized using a least square minimizing algorithm from an inner routine in software Mathematica (see [11] for algorithm description). This set of parameters is used in section 3 to adjust the age-structured model 3.1. The values for the parameters were found to be

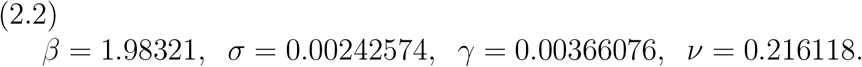

The initial conditions used for the algorithm were *β*^∗^ = 0.8481, *σ*^∗^ = 0.2682, *γ*^∗^ = 0.0870, which were obtained from the national case (see [3]).

Figure 1 shows the real data and the SEIR curve of infected individuals using the parameters from (2.2). The time interval considered was 15 days.

**FIGURE 1.**
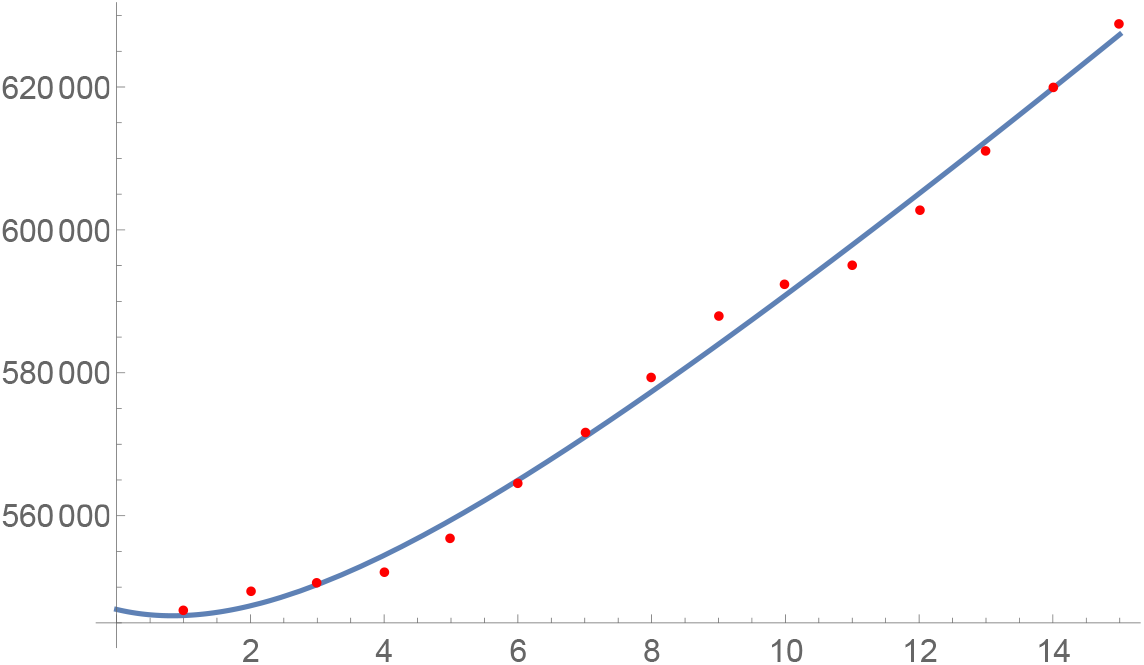
The fitted curve for the total number of infected individuals for the classical SEIR model together with the real data.

The effect of the vaccination on the prevalence curve, *I*(*t*), is twofold: it decreases the maximum value of it but it anticipates the date of its occurrence, see figure 2.

**FIGURE 2.**
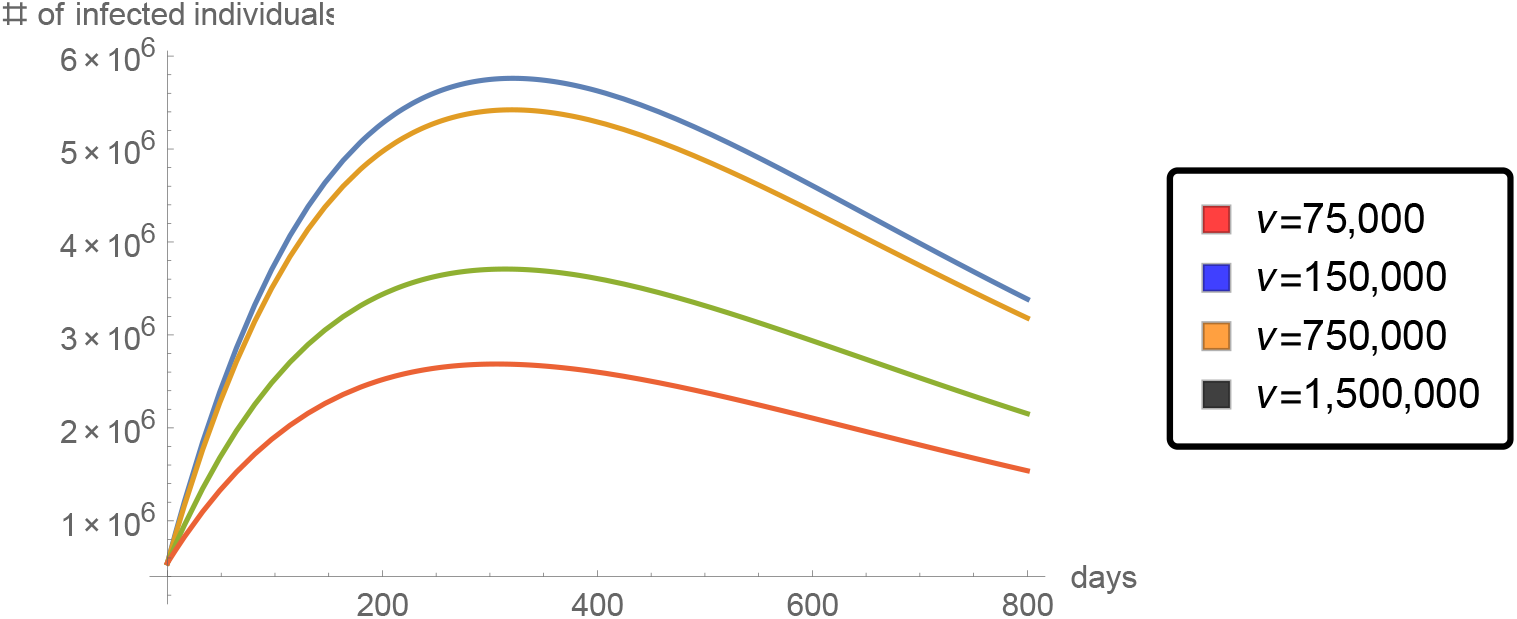
Total number of infected individuals for several values of daily effort of vaccination *ν*.

This reasoning could lead us to ask “how much earlier is the Public Health System of Minas Gerais (PHS-MG) able to deal with such a maximum of the number of infected cases ?” In fact we need not to worry about this question because we observe that, the summit reached with the vaccination strategy going on, is lower than the corresponding value for the number of infected individual if no vaccination was taking place. Yet we want the PHS-MG to be able to deal with such a high number of infected individual in need of intensive care units (ICU).

According to the its health department (see [5]) the state of Minas Gerais has 4, 199 ICU beds in the PHS-MG, so that 3, 360 corresponds to 80% of this total and we shall analyze what should be the vaccination effort, *ν* necessary to keep the level of occupation of such beds below this number. According to BBC web site (see [2]) between 5% to 15% of the total number of infected individuals needs intensive hospital care. This implies that the maximum number of simultaneously infected individuals in Minas Gerais should be between 22, 400 and 67, 200. The average period of hospitalizations is accounted to be around 15 days. So, we consider the function *inf* (*ν, t*) and *rem*(*ν, t*) representing the total number of infected individuals and the total number of removed (causalities+recoveries) individuals respectively, numerically obtained from the system of differential equations given by (2.1) and we construct the auxiliary function of simultaneously infected individuals as *sick*(*ν, t*) = *inf* (*ν, t*) − *rem*(*ν, t*) − (*inf* (*ν, t* − 15) − *rem*(*ν, t* − 15)) the graphic of which, for several values of the vaccination parameter, *ν*, is presented in figure 3 together with the maximum accepted numbers of simultaneously infected individuals *L*1 = 22, 400 and *L*2 = 67, 200.

**FIGURE 3.**
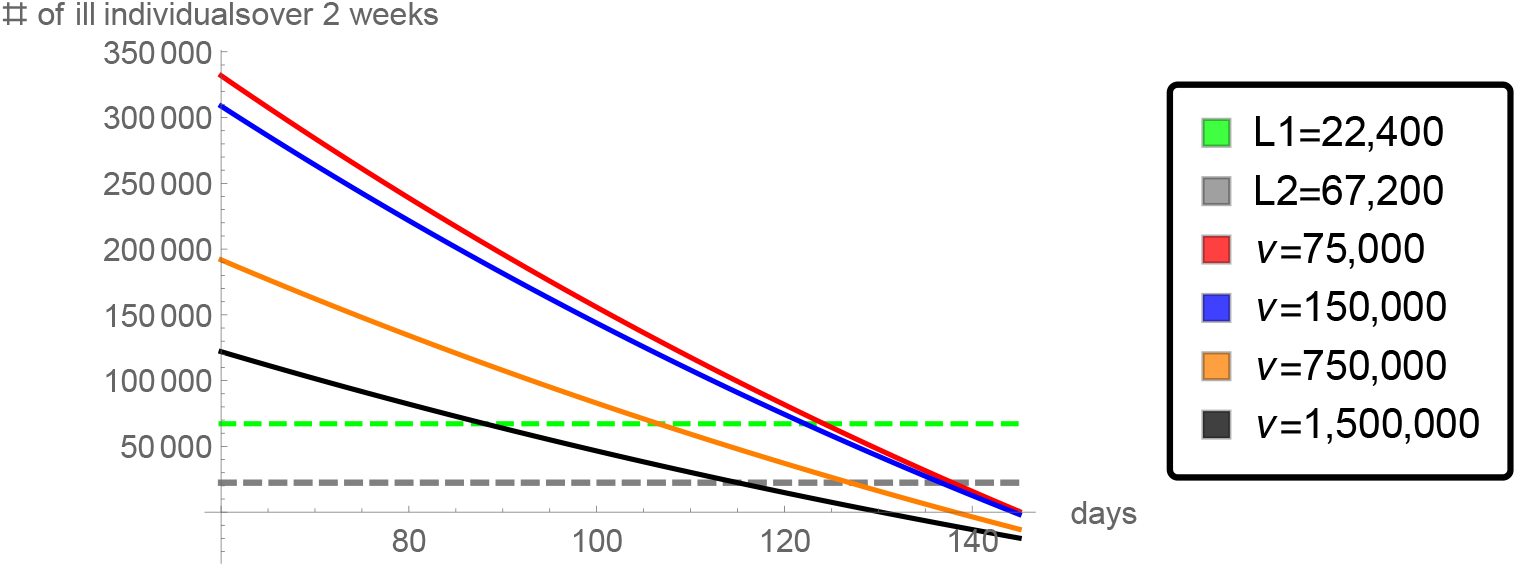
Total number of ill individuals in 2 week-time.

This result indicates that even if we drastically increase the current rate of number of vaccinated individuals per day from 75, 000 to 1, 500, 000 the number of individuals simultaneously in need of an ICU bed would be below 80% of the full capacity of the PHS-MG around 90 days (or 120 days in the worst scenario) after the effects of the vaccination has begun.

## 3. The Age-structured SEIR model

Since age is an important factor on the COVID-19 epidemic, it will be assumed that the population is age structured (see [6], [14], [4] for continuous models and [10], [15] for discrete models). Three age classes are used; *i* = 1 : infants with ages in the interval [0, 17], *i* = 2 : adults with ages in the interval [18, 59], and *i* = 3 : elderly with ages in the interval [60, 110].

Let *S*_*i*_(*t*), *E*_*i*_(*t*), *I*_*i*_(*t*), *R*_*i*_(*t*) represent the number of susceptible, exposed, infected and removed individuals at age class *i* respectively at time *t* ≥ 0 measured in days. The system of ordinary differential equation we wish to analyze is

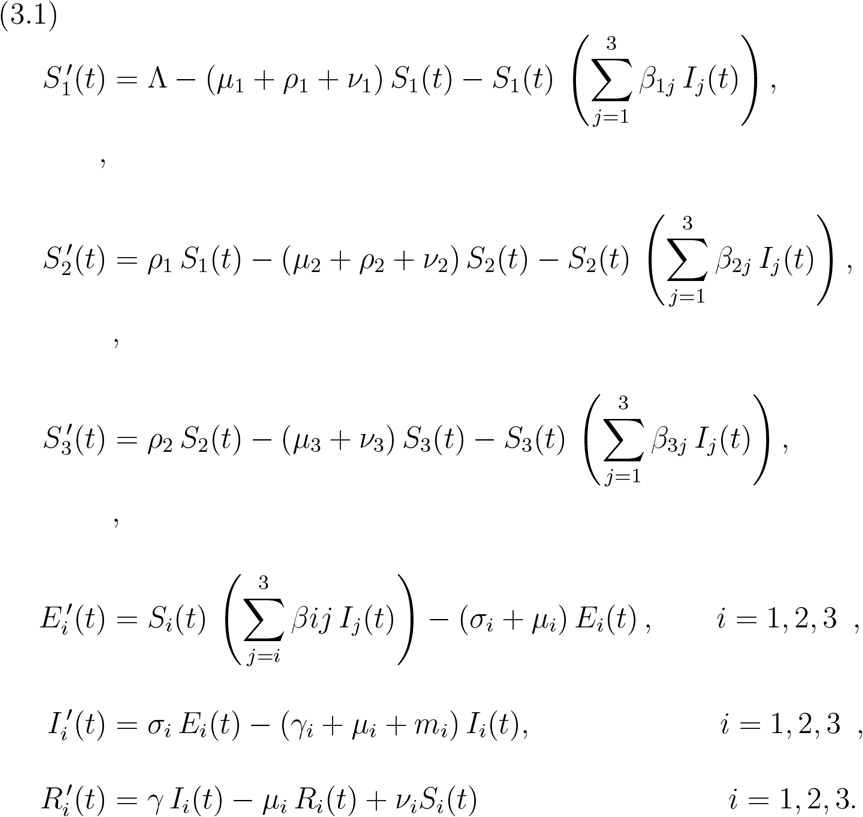

The parameters are all non-negative and they are described in Table 1. Typically, it will be assumed that *β*_*ij*_ = *β*_*ji*_.

**TABLE 1.** Parameters of the basic SEIR model with vital dynamics.

| Parameter | Description |
| --- | --- |
| $\nu_i$ | vaccination rate for class $i$. |
| $\Lambda$ | recruitment rate. |
| $\mu_i$ | natural death rate for class $i$. |
| $\rho_i$ | survival rate for class $i$ to class $i + 1$ $i \leq 2$. |
| $\beta_{ij}$ | pathogen's transmission rate between classes $i$ and $j$. |
| $\sigma_i$ | rate at which exposed of class $i$ convert into the infected class. |
| $\gamma_i$ | class $i$ host's recovery rate. |
| $m_i$ | host's pathogen-induced death rate at class $i$. |

The total population of the state of Minas Gerais is of 21, 204, 000 inhabitants at the end of 2019 and the age distribution is given in table (2), (see [8])

According to the daily epidemiological bulletin of the health department of January 1^*st*^, 2021 Minas Gerais has had 546, 884 confirmed cases of COVID-19 and a total number of 12, 001 deaths officially caused by the disease featuring a lethality rate of *η* = 2.2%.

According to [8] Minas Gerais had 258, 665 births and 141, 451 deaths in 2019, featuring a net increase of 0.55% by the end of 2019. We assume the casualties caused by Covid are totally atypical so we estimate the total population of Minas Gerais at the end of 2020 to be 1.0055 × 21, 204, 000 − 12001 = 21, 308, 621. According to health department of the State of Minas Gerais (see [8]) on January 1^*st*^, 2021 we had the total number of infected, exposed and removed individuals given by, *I*_0_ = 546, 884, *E*_0_ = 40, 605 and *R*_0_ = 506, 279 respectively so that we can consider the total number of susceptible individuals to be *S*_0_ = 21, 308, 621 − (*I*_0_ + *E*_0_ + *R*_0_) = 20, 214, 853.

Let *N* denote the total population and *D* the total deaths per year, thus 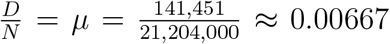. Let *D*_*i*_ and *N*_*i*_ be the number of deaths per year and *N*_*i*_ be the population of age class *i* respectively. Thus 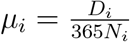. With this notation the data on Table 2 is denoted by

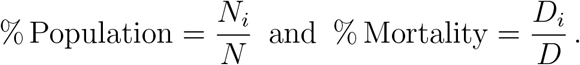

Since *µ*_*i*_ must be considered in a daily base it is calculated by

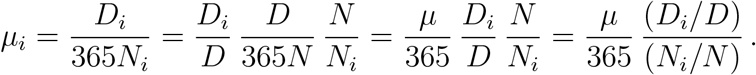

**TABLE 2.** Age Classes.

| Class | Age (years) | % Population ($c_i$) | % Mortality (year) | $\mu_i$ (day) |
| --- | --- | --- | --- | --- |
| 1 | [0,17] | 22.49 % | 3.68 % | $3 * 10^{-6}$ |
| 2 | [18,59] | 60.22 % | 25.4 % | $7.7 * 10^{-6}$ |
| 3 | [60,100] | 17.29 % | 70.92 % | $7.5 * 10^{-5}$ |

As already mentioned, we analyze the development of the epidemic from January 1^*th*^, 2021 onwards, therefore, by allusion to the usual situation, we shall call it the “steady state”. We denote by 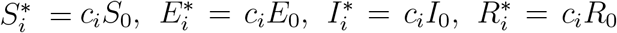 and at the steady state we have

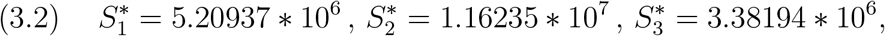

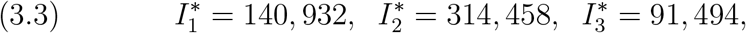

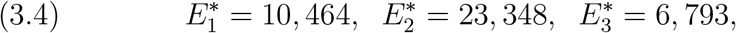

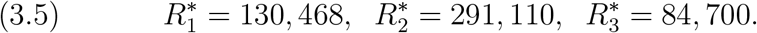

Adding the equations for the steady state features

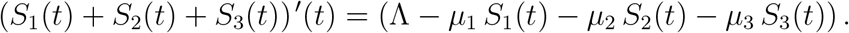

Assuming that the total population is constant and on demographic equilibrium, using the values for the population distribution at the equilibrium values, one must have

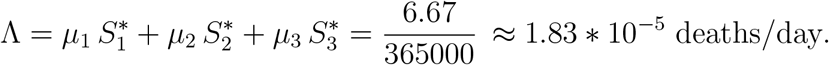

Furthermore, once the time interval of interest is small compared to the demographic time scale, the actual annual growth rate will not be taken into consideration. The demographic equilibrium implies that *ρ*_1_ and *ρ*_2_ satisfy

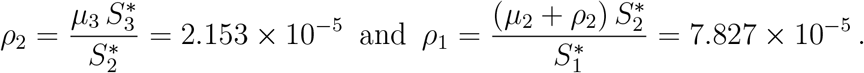

We shall also assume the demographic and disease parameters are the same for all age classes.

The values of the mortality rate induced by the COVID-19 are obtained from the epidemiologic bulletin of the state health department of Minas Gerais of day one, i.e. January 1^*St*^, 2021 (see [5]) and is presented in table (3).

**TABLE 3.** Death rates for the age structured model.

| Age group | Number of cases | Deaths | $m_i$ |
| --- | --- | --- | --- |
| 1 | 76671 | 45 | 0.06% |
| 2 | 706130 | 3964 | 0.56% |
| 3 | 164701 | 16072 | 9.7% |

## 4. Control vaccination strategies for the age-structured model

Before we proceed, we need to adjust the parameters for the age structured model. Throughout this analysis we suppose the vaccination does have an impact on the level of transmissibility of the virus. This is a reasonable supposition not yet proved but taken for granted in this study.

### 4.1. Data Fitting

There are 15 parameters to be determined for the age structured model:

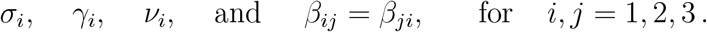

The algorithm fits these parameters to the available data of Minas Gerais’s total number of reported cases for the number of infected individuals of the 15 first days of 2021 by a least squares method. The distance between the predicted curve

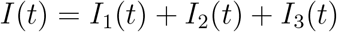

and the data curve is minimized. The initial parameters for the minimization search algorithm are the ones found for the classical (unstructured) SEIR model (2.1) taking into consideration the population percentage of each age class. Let *c*_*i*_ be the population percentage of each class, that is (see Table 2), *c*_1_ = 0.2249, *c*_2_ = 0.6022 and *c*_3_ = 0.1729. The initial values for the interaction are chosen as

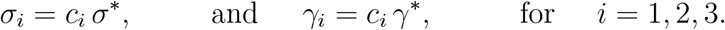

and

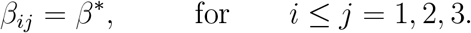

The resulting values are listed in Table 4.

**TABLE 4.** Fitted parameters for the age-structured SEIR model without vital dynamics.

| $\beta_{ij}$ | Value | $\beta_{ij}$ | Value | $\sigma_i$ | Value | $\gamma_i$ | Value | $\nu_i$ | Value |
| --- | --- | --- | --- | --- | --- | --- | --- | --- | --- |
| $\beta_{11}$ | 1.79061 | $\beta_{22}$ | 0.65525 | $\sigma_1$ | 0.0543033 | $\gamma_1$ | 0.386621 | $\nu_1$ | 0.205256 |
| $\beta_{12}$ | 0.377533 | $\beta_{23}$ | 0.256922 | $\sigma_2$ | 0.144925 | $\gamma_2$ | 0.130522 | $\nu_2$ | 0.213013 |
| $\beta_{13}$ | 1.3625 | $\beta_{33}$ | 0.649881 | $\sigma_3$ | 0.126595 | $\gamma_3$ | 0.155331 | $\nu_3$ | 0.211383 |

The control measures for the age structured model with vaccination will be done through a numerical analysis of the total number of infected individuals when different vaccination strategies are used. Although none of the vaccines available at this moment has shown any study of real effectiveness when used in the youngsters we shall present our numerical simulations of such a possible vaccination strategy for it could be shown to be useful in the future.

Firstly we compare the strategy of vaccination involving only the youngster and the adults. In Figure 4 (left) we show the total number of infected individuals when the full effort of vaccination is put on vaccinating either only the youngster or only the adults. We clearly see that the strategy of focusing the vaccination totally on the youngsters does not produce a high decrease in the total number of infected individuals as the strategy of focusing the vaccination totally on the adults. Similarly in Figure 4 (right) we show the total number of infected individuals when the full effort of vaccination is put on vaccinating either only the youngster or only the elderly. We clearly see that the strategy of focusing the vaccination totally on the youngsters does not produce a high decrease in the total number of infected individuals as the strategy of focusing the vaccination totally on the elderly. Clearly a deeper study would be necessary to really determine whether another choice of strategy involving the youngster could produce higher decrease in the total number of infected individuals. We prefer to let this analysis for a future study and proceed here with a focus on the comparison between vaccination strategies involving only the age groups of adults and elderly.

**FIGURE 4.**
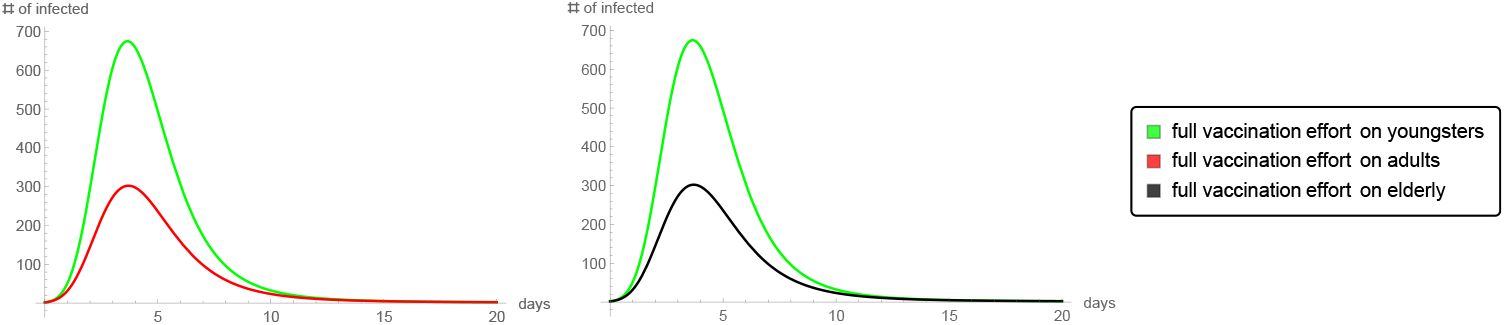
The total number of infected individuals when only youngster or only adults are vaccinated (left). The total number of infected individuals when only youngster or only elderly are vaccinated (right).

The immunization strategy used in the state of Minas Gerais follows the national plan of focusing the vaccination on the more vulnerable people which, due to its high lethality rate, clearly is the age group of elderly. We study here whether this strategy is really a good one. Minas Gerais started vaccinating its population on January 25^*th*^. Due to the lack of sufficient quantities of vaccine doses by April 9^*th*^ only around 10% of the population of Minas Gerais had been vaccinated and the great majority of it were the elderly (the government vaccination policy prioritizes the elderly and the health agents working on the front lines). By April 9^*th*^ around 70, 000 people/day were being vaccinated. We analyze the total number of infected individuals in Minas Gerais in all possible ways of distributing the daily 70, 000 doses of vaccines among adult and elderly individuals only. In order to do that we consider a function, *F*, of the total number of infected individual of ages groups 2 (adults) and 3 (elderly) at time *t* considering a daily rate of vaccination *ν*_*i*_ for the age class group *i*. So we have *F* (*t, ν*_2_, *ν*_3_) and then we consider the daily rate (of beginning of April) of individuals been vaccinated in Minas Gerais to be *V max* = 70, 000. We have analyzed the function *F* (*t, rV max*, (1 − *r*)*V max*) for all values of *t* ∈ [0, 60] and *r* ∈ [0, 1] which clearly represents the effects over the total number of infected individuals during 60 days after the vaccines start to work and considering all the possible ways of distributing the vaccines among adults and elderly. Because the values of function F are very big and its variation is very small, in order to have a better visualization of the results, we prefer to shows the graphic of an auxiliary function: *G*(*t, r*) = *Exp*(*F* (*t, rV max*, (1 − *r*)*V max*)*/*10^6^) together with the graphic of the plane *z* = 3.5 in such a way that we can more clearly see that the minimum does occurs at *r* = 1 (see figure (5)). Figure (6) shows the graphics of *G* when *r* = 0, *r* = 0.25, *r* = 0.5, *r* = 0.75 and when *r* = 1. We can clearly see that at a short time interval (20 days after the effects of the vaccines begin to work) the minimum number of infected individuals is obtained when *r* = 1, that is to say when all the vaccination effort is put on the age group of the adults. For a longer time interval the graphics point out that the minimum occurs when *r* = 0.

**FIGURE 5.**
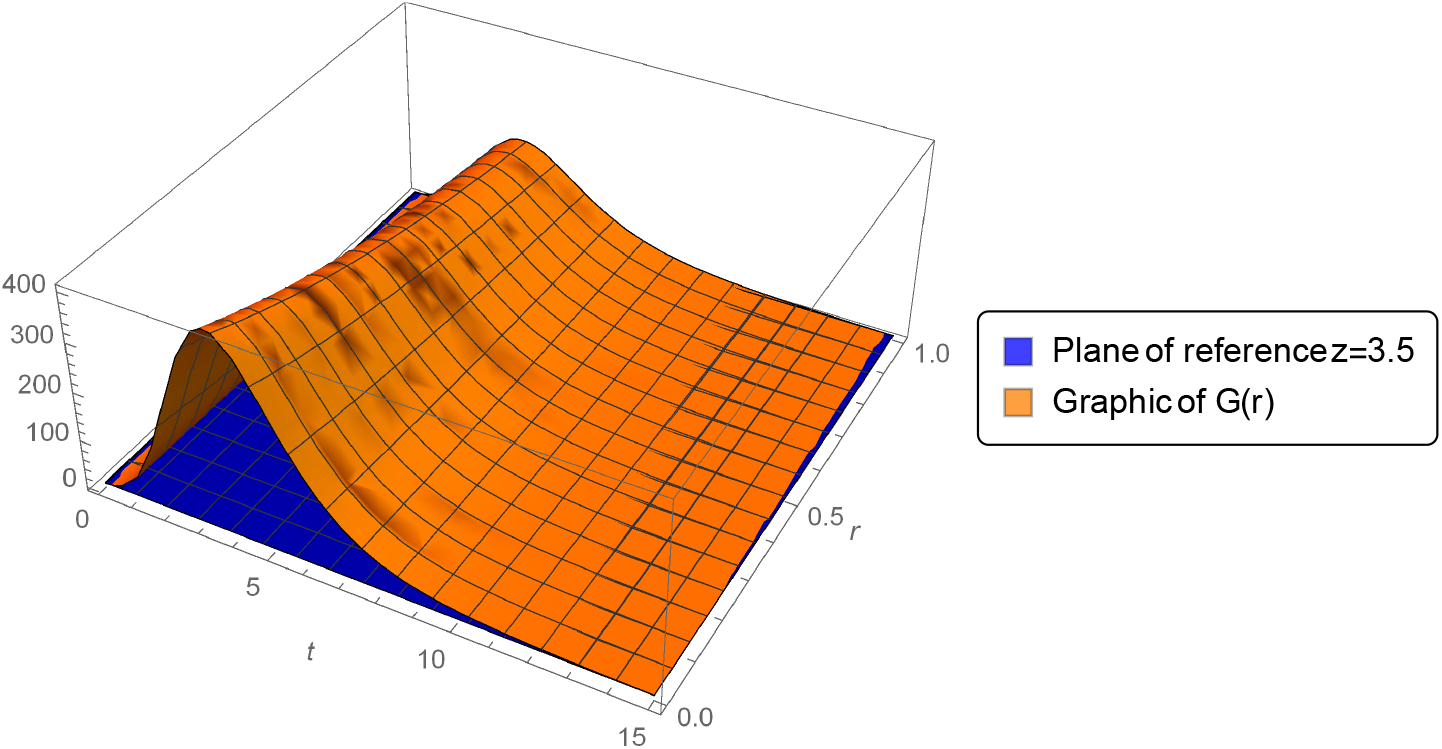
The graphic of *G*(*t, r*) and a plane of reference as described in the text for the time interval *t* = 0 till *t* = 15.

**FIGURE 6.**
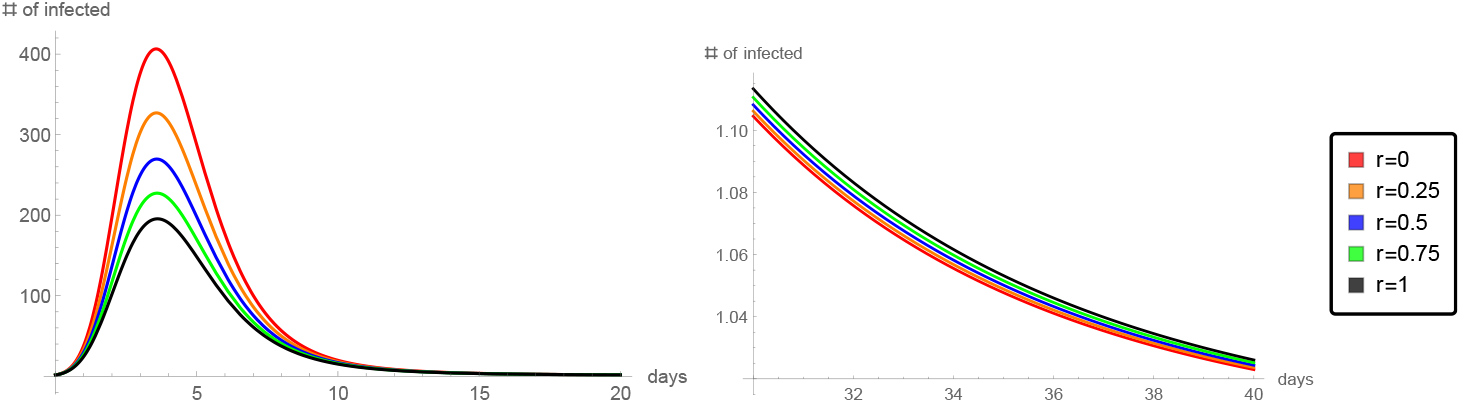
The graphic of *G*(*r*) for the time interval *t* = 0 to *t* = 20(left) and from *t* = 30 to *t* = 40 (right).

## 5. Conclusions

We have introduced an age structured SEIR model with vaccination. Three age groups were considered: infants (0 to 17 years), adults (18 to 59 years) and elderly (60 to 110 years). Firstly we studied the associated classical (unstructured) SEIR model without vital dynamics. The parameters were fitted by a least-square algorithm and the impact of the vaccination parameter *ν* was studied. As we expected the maximum number of infected individuals decreases as the vaccination rate increases. Also it was observed that the maximum number of infected cases happens earlier than if no vaccination were done, yet such a pick is still smaller than the corresponding value of the number of infected individuals of the non vaccination situation which clearly justifies the strategy of vaccination at any rate.

However it has also been shown that at the current rate of vaccination or even if such rate is multiplied by 20, vaccination procedures alone are not enough to keep the number of seriously ill hospitalized individuals below 80% of the public health system full capacity. Thus, this study clearly indicates that the need of strict rules of social distancing, frequent hands washing and other similar measures of avoiding crowdedness should be implemented and stimulated by the local governments.

Furthermore, in order to analyze different vaccination strategies the parameters obtained for the unstructured SEIR model were used to adjust the parameters for the age-structured SEIR model with vaccination. We compared vaccination strategies involving only adults and elderly for this case is closer to the present reality since at the moment non of the vaccines under use in the world has been completely tested on youngsters. We postpone to future works the studying of different strategies of vaccination involving youngsters. Using this data, we analyzed the possible strategy of vaccinating *rV max* adults and (1 – *r*)*V max* elderly, letting *r* ∈ [0, 1]. We have numerically computed the total number of infected individuals and have shown that the strategy with *r* = 1 produces the best results. The final conclusion is that in order to minimize the number of infected individuals the current strategy of vaccination with total focus on the elderly (*r* = 0) has to be radically changed to a vaccination strategy with total focusing on the adults, i.e. only adults should be vaccinated (at least at the first 20 days) if the current rate of *V max* = 70, 000 individuals per day is maintained.

## Data Availability

All the data used in the article has been referred to at the bilbiography and can be easily found on the corresponding website

## Acknowledgement

Not applicable

## Footnotes

1 World Health Organization Website https://www.who.int/

